# A Systematic Review of Female Athletes in Concussion Research: Gaps, Findings, and Future Directions

**DOI:** 10.1101/2025.04.15.25325892

**Authors:** Sterling Gutterman, Rachel Edelstein, John Darrell Van Horn

## Abstract

**Background:** Despite the growing participation of female athletes in contact and high-impact sports, their representation in concussion research remains disproportionately low. This imbalance limits the generalizability of clinical guidelines and return-to-play (RTP) protocols, potentially compromising athlete safety and recovery outcomes.

**Methods:** A systematic literature review was conducted on concussion studies published between 2021 and 2024 to assess changes in female representation. The analysis focused on three key objectives: (i) identifying primary research areas in concussion studies, (ii) evaluating the prevalence of female participants, and (iii) determining whether sample sizes were sufficient for statistically meaningful, sex-specific conclusions.

**Results:** A regression analysis revealed a statistically significant decline in female representation over time (B =-0.2478, p <.001), and female participant numbers decreasing 25% yearly and by 31% between 2021 and 2024. The female-to-male ratio analysis confirmed that male sample sizes continue to dominate, reinforcing the reliance on male-centric data in concussion assessment and RTP decision-making. Additionally, while generalized linear models remain widely used for broad comparisons, traditional statistical methods often fail to capture nuanced sex-based differences, particularly given the persistent sample size disparities.

**Conclusion:** This review underscores the continued underrepresentation of female athletes in concussion research and the reliance on male-driven data for clinical decision-making. Although efforts to include female participants have increased, sample sizes remain insufficient for robust, sex-specific analyses. Addressing these gaps is critical for developing evidence-based, individualized concussion recovery models that account for the unique neurobiological and biomechanical profiles of female athletes.

## Introduction

Historically, concussion research has primarily focused on male athletes, leading to a significant gap in understanding how concussions uniquely affect female athletes (D’Lauro et al., 2020; Edelstein et al., 2024). Despite women’s increasing participation in contact and high-impact sports, their representation in concussion studies remains disproportionately low, limiting the generalizability of clinical guidelines and return-to-play protocols (Covassin et al., 2013).

Emerging research suggests that female athletes experience distinct and often more severe outcomes following SRC compared to males (Bretzin et al., 2022; Mollayeva et al., 2018). Biological and biomechanical differences contribute to these disparities, including but not limited to variations in brain structure, hormonal fluctuations, and musculoskeletal factors (Master et al., 2021; Wunderle et al., 2014). For instance, female axons tend to have smaller diameters and reduced white matter density, making them more susceptible to shear forces and diffuse axonal injury following a concussive event (Master et al., 2021; Taylor et al., 2022).

Additionally, neuroimaging studies suggest differences in functional brain connectivity between male and female athletes, further influencing concussion symptom severity and recovery time (Vedung et al., 2022).

Another significant factor contributing to sex-based differences in concussion outcomes is neck musculature and biomechanics. Female athletes typically have less neck strength and muscle mass than male athletes, leading to rapid head acceleration upon impact, which increases concussion risk and injury severity (Lynall et al., 2019). Research has demonstrated that stronger neck musculature may reduce concussion risk by dissipating impact forces, suggesting a potential avenue for targeted prevention strategies (Mihalik et al., 2016). For instance, a study by Raftery et al. (2020) highlighted that increased neck strength can absorb and distribute forces during collisions, further emphasizing the protective role of neck strength and unique treatment in reducing concussion risks. Additionally, Kowalewski et al. (2021) found that training protocols focused on enhancing neck strength in female athletes significantly decreased the incidence of concussions in high-contact sports. These findings underline the importance of integrating neck strengthening exercises into training regimens to potentially mitigate concussion risks among female athletes.

Hormonal fluctuations further complicate concussion recovery in female athletes. Studies indicate that the menstrual cycle phase during injury can significantly impact symptom severity and recovery duration (Duffy et al., 2021; Wunderle et al., 2014). Concussions sustained during the luteal phase—characterized by elevated progesterone levels—are associated with prolonged symptoms and delayed recovery (Bazarian et al., 2013). A study published in the Journal of Head Trauma Rehabilitation found that women who experienced mild traumatic brain injuries (mTBIs) during the luteal phase reported poorer health outcomes and slower recovery one-month post-injury compared to those injured during the follicular phase or those on hormonal contraceptives (Bazarian et al., 2014).

Concussions may involve a sudden drop in progesterone levels following the injury, leading to a “withdrawal” effect during the luteal phase, suggesting more severe and prolonged symptoms (Bazarian et al., 2014; Natural Womanhood, 2020). Additionally, hormonal fluctuations post-injury can disrupt the hypothalamic-pituitary-ovarian axis, potentially leading to menstrual irregularities and further complicating recovery (Concussion Treatment Hub, 2021). These findings underscore the importance of considering menstrual cycle phases when assessing and managing concussions in female athletes (Bretzin et al., 2022; Hardaker et al., 2024).

Beyond these physiological differences, clinical research has shown that female athletes report higher symptom severity across multiple domains, including cognitive (e.g., memory deficits, brain fog), vestibular (e.g., dizziness, balance issues), and emotional (e.g., anxiety, depression) symptoms (Covassin et al., 2013; Alla et al., 2009; Brown et al., 2015). Additionally, female athletes tend to experience prolonged recovery times, with higher rates of post-concussion syndrome (PCS), where symptoms persist beyond four weeks (Master et al., 2021; McCrea, Broglio, & McAllister, 2021). This extended recovery period increases the risk of repeat concussions, which can lead to cumulative brain damage and long-term cognitive impairment (Zuckerman et al., 2015).

Despite growing evidence of sex-based differences in concussion recovery, female representation in concussion studies remains disproportionately low, making it difficult to draw statistically significant conclusions and develop sex-specific clinical guidelines (Edelstein et al., 2024; McCrea et al., 2005). Many standardized return-to-play (RTP) protocols and symptom assessment tools, such as the SCAT symptom checklist, were developed using predominantly male samples, limiting their applicability to female athletes (Broshek et al., 2005; D’Lauro et al., 2022; Edelstein et al., 2023). To address these limitations, researchers advocate for individualized recovery models that consider sex-specific symptom profiles, hormonal influences, and biomechanical vulnerabilities. Future research should prioritize expanding female athlete cohorts in concussion studies to improve the generalizability of findings.

Based on the D’Lauro et al. (2022) systematic review, which showed 80% of concussion research up until 2020 is male dominant, with 40% of those samples including zero female participants, a non-exhaustive review of literature from 2021-2024 was done to see if trends have changed over the last five year. The key goals are as follows: (i) show what areas of the field are researched, (ii) define the prevalence of female participants, and (iii) determine if the number of female participants produces results with enough significant power. Thus, we hope to raise awareness about the current gaps in research and provide evidence for why studying females differs from males. Despite increasing female inclusion in current research, we hypothesize that the quantity is too low to produce significant results, highlighting the need for further research.

## Methods

To collect data on the current landscape of research regarding the unique effects of concussions on female athletes, it is essential to conduct a thorough literature review. This collection examines publications from the past three years (2021–2024) to complement previous research on trends before 2021. PubMed served as the primary database for sourcing relevant studies, with key search terms including various combinations of “female,” “sport-related injury,” “return-to-play,” and “neuroimaging.” Additionally, reference sections, as well as PubMed’s “cited by” and “similar articles” features, were used to identify additional studies that aligned with the scope of this review.

While individual experimental studies were prioritized, a wealth of meta-analyses also provided valuable summaries on sport-related concussions and their unique impacts on female athletes. It was also essential to include studies examining sports played by both males and females and to ensure that the research was conducted within a year of the article’s publication. Infromation on the selected studies is as follows: the reviewed studies were published in 2021 (6), 2022 (2), 2023 (4), and 2024 (3); research was conducted globally, with studies originating from the United States (9), Australia (2), Canada (2), England (1), and Sweden (1); these studies were published in a range of academic journals, including *JAMA Neurology* (2), *Journal of Neurotrauma* (2), *Human Brain Mapping* (2), *British Journal of Sports Medicine* (2), *Journal of Alzheimer’s Disease Reports* (1), *Journal of Neurology* (1), *Sports Medicine* (1), *Brain Injury* (1), *BMJ Open Sport & Exercise Medicine* (1), *Journal of Clinical and Experimental Neuropsychology* (1), and *Sleep Health* (1).

Several key factors were identified in the research, including the year of publication and authorship, which helped determine source credibility. Other considerations included total sample size to ensure robust results, the breakdown of male and female subjects to assess representation, and the statistical methods used for analysis. First, we calculated statistical summaries of various variables, including total sum, mean, median, maximum, and minimum values. Next, we focused on sample size and conducted various power analyses to determine the required number of male and female participants in each study to achieve proper statistical power and significance. Power analyses were performed in R (R Core Team, 2025). Subsequently, we visualized sample sizes and attempted to establish a regression line to understand trends in participation over four years.

Lastly, we investigated the statistical methods used in the studies. The research spanned biomarkers, blood flow alterations, brain microstructure, somatic symptoms, and recovery metrics, each of which may align better with certain methods. Commonly employed statistical methods included descriptive statistics, ANOVA calculations, linear and logistic regressions, and partial least squares (PLS). To identify these results, we searched each article’s specific statistical methods section for different methods, documenting those that analyzed similar variables.

To analyze trends in sample representation and statistical methodologies, the methods section of each article was reviewed to extract relevant data. A linear and logistic regression analysis were conducted to examine trends in female representation over time, where the year of publication served as the independent variable and the female sample size as the dependent variable. Additionally, statistical methods reported in each study were documented and categorized based on key terms to identify common analytical approaches.

A comprehensive power analysis determined the adequate sample size required to sustain a meaningful and statistically significant result. This process involved evaluating various factors that influence sample size, including the expected effect size, the desired power level, and the significance level, to ensure that the study would be adequately equipped to detect actual effects if they exist. By identifying the optimal sample size, the analysis aimed to enhance the reliability of the findings and contribute to the robustness of the research outcomes.

To conclude the review, the frequency of each statistical method employed was meticulously quantified.

A pie chart was produced to visually represent these findings, illustrating the proportion of each statistical method in relation to the total number of techniques utilized across all the studies considered. This systematic approach offered a clear and structured overview of the prevailing methodological trends and highlighted the level of female representation within concussion research. By analyzing these statistical methods and assessing their distribution, we aimed to illuminate potential biases or gaps within the research landscape, ensuring a more holistic understanding of how gender dynamics influence this field of study.

### Lack of Concussion Studies within 2021-2025

Despite a rigorous and thorough search process employing various academic databases and research methods, 37 scholarly articles published between 2021 and 2025 were identified as relevant to the topic at hand.

While more studies were evaluated, finding harmony between year, country, sample size, and sport presented significant challenges. It is important to note that there is a noticeable gap in the currently available completed research on this topic, highlighting the need for more extensive investigation into the nuances and complexities associated with the subject matter. Consequently, the subsequent sections of this work will present a comprehensive literature review that critically examines the fundamental factors contributing to the findings within this area of study. This review aims not only to provide a thorough understanding of the implications of gender in sports-related concussion research, but also to highlight the significant areas where further inquiry is warranted. By addressing these shortcomings, the literature review intends to foster a deeper comprehension of the sociocultural dynamics at play and encourage future research endeavors that can contribute to a more complete understanding of the issue.

## Results

A total of 37 articles were screened for this review, with N=14 articles meeting the following inclusion criteria: (i) Study conducted between 2021-2025, (ii) contained both male and female samples, (iii) specified sample sizes. The results of this literature review were then analyzed based on several article characteristics, including publication journal, publication year, number of authors, host country, sample size, and statistical analyses used. Regarding author count, 172 authors contributed to the reviewed articles, with an average of 11 authors per study. The number of authors can serve as an indicator of interest in the topic within the field, the diversity of perspectives represented, and the overall accuracy and rigor of the research.

Across the 14 selected studies, 4,075 participants were included, comprising 2,380 males and 1,695 females (41.60%). Figure 1 illustrates the variation in female athlete inclusion over the past four years and compares it to male representation. The linear regression analysis examining the relationship between year of publication and female sample size revealed a negative trend in female representation in concussion research. The regression equation, Female Sample Size =-31.44 × Year + 63702.36, indicates that the predicted female sample size decreases by approximately 31 participants for every one-year increase. This downward trajectory suggests that, despite increasing awareness of sex-based differences in concussion outcomes, the inclusion of female participants in research studies has not improved and is declining over time. The negative slope reinforces the concern that current recruitment strategies may not be prioritizing gender balance, ultimately limiting the generalizability of concussion findings to female athletes. If this trend continues, future studies will lack sufficient statistical power to analyze sex-specific concussion recovery patterns, emphasizing the urgent need for targeted interventions to increase female participation in concussion research.

**Figure 1.**
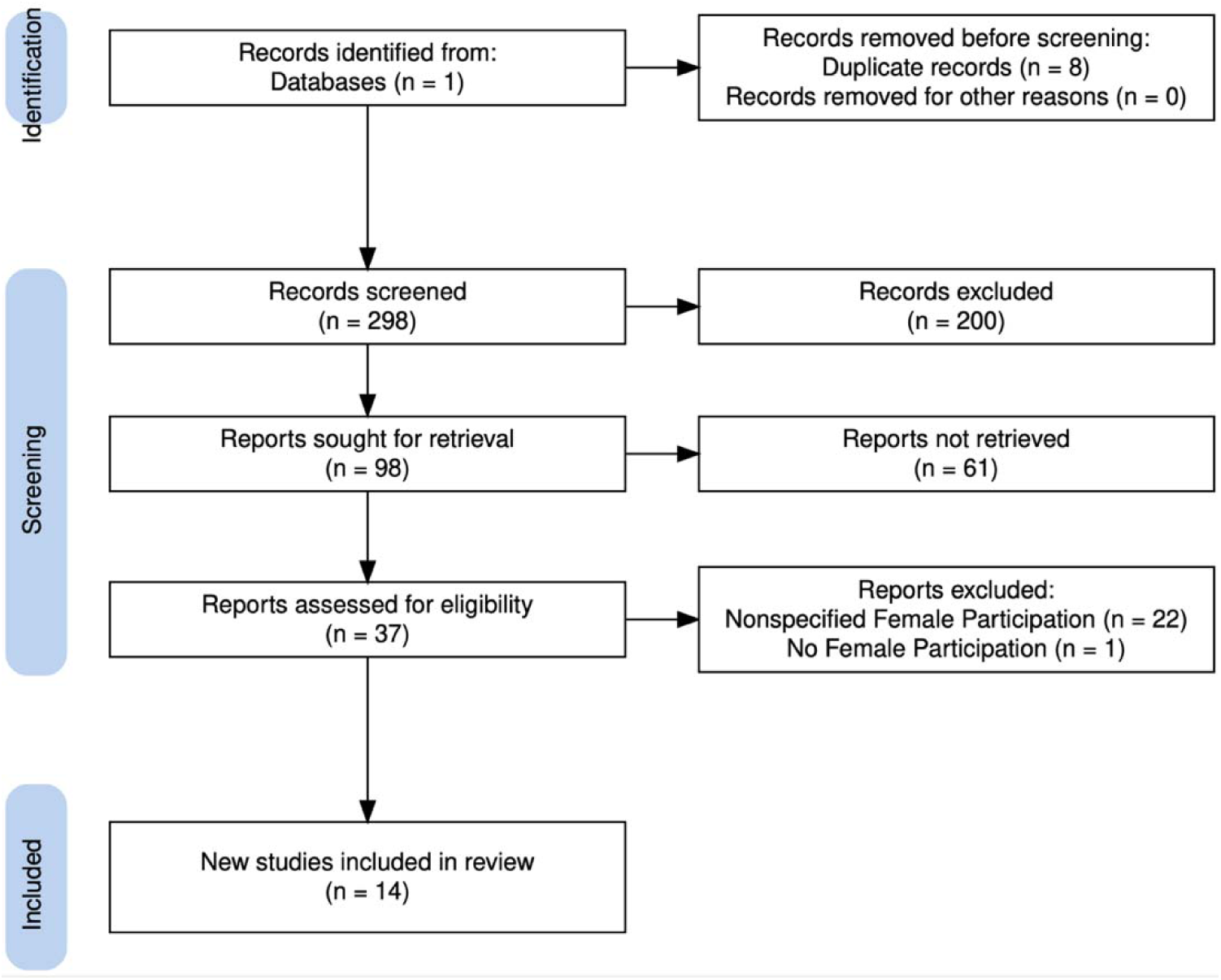
PRISMA flow diagram illustrating the study selection process for the literature review. A total of 298 records were screened after removing duplicates. Following title and abstract review, 98 reports were sought for retrieval, of which 37 were assessed for eligibility. Ultimately, 14 studies were included in the final review. The most common reasons for exclusion at the eligibility stage were unspecified female participation (n = 22) and lack of female participants (n = 1), underscoring the gender representation gap in concussion research.

Figure 2 presents the linear regression results of sample sizes by gender across journal articles published between 2021 and 2024 in concussion research. The bar chart distinguishes female (red) and male (orange) sample sizes, with the y-axis representing sample size and the x-axis representing publication year. The figure presents sample sizes by gender across journal articles published between 2021 and 2024 in concussion research. The bar chart differentiates between female (red) and male (orange) sample sizes, with the y-axis representing sample size and the x-axis representing the year of publication. Across all years, male sample sizes consistently exceed female sample sizes, highlighting a persistent gender disparity in concussion research.

**Figure 2.**
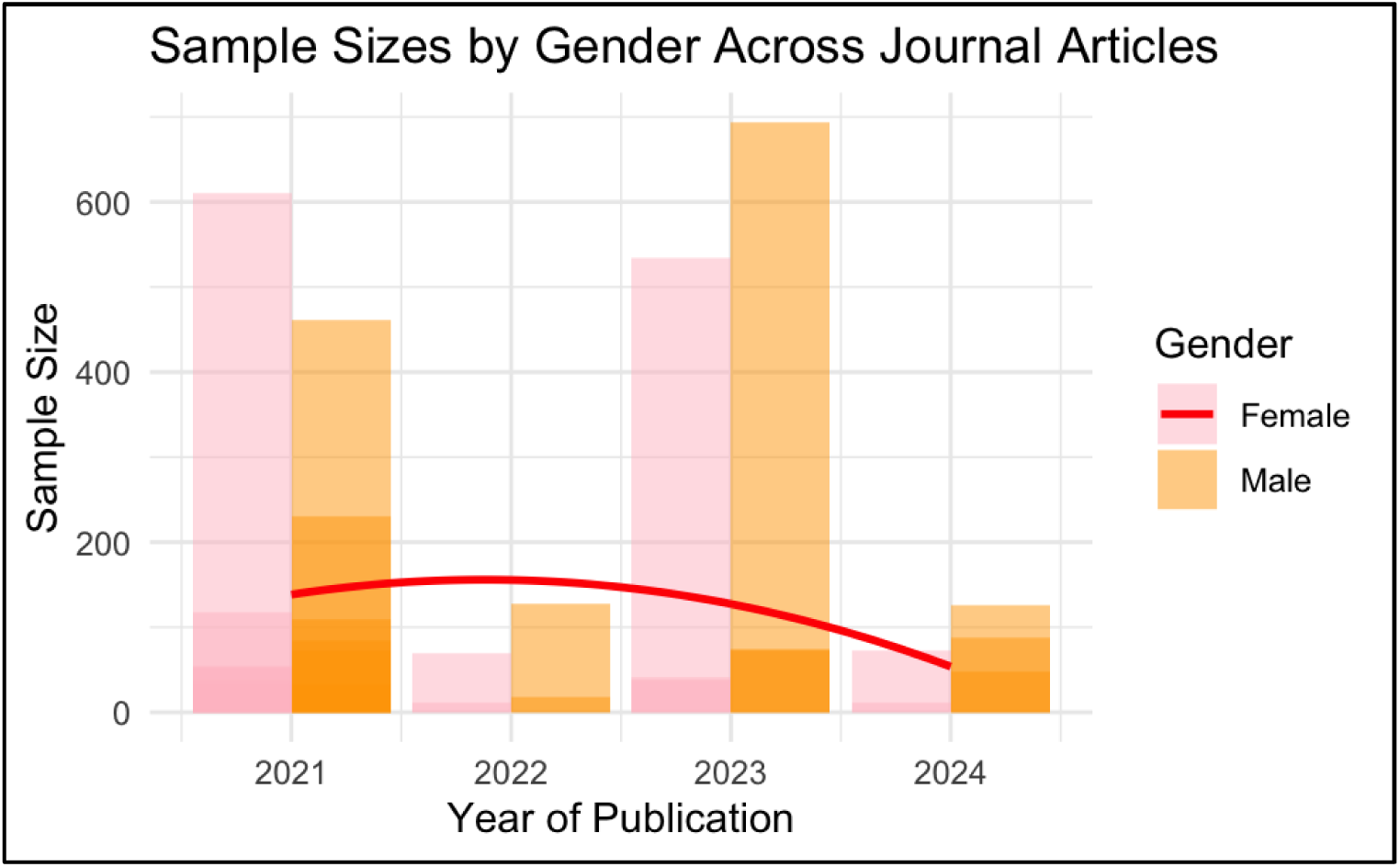
Frequency of Sample Sizes by Gender and Year. Sample sizes by gender across journal articles published between 2021 and 2024. Bars represent the cumulative number of male (orange) and female (pink) participants included in concussion-related studies for each year. While female representation approached parity in 2021, subsequent years revealed a consistent underrepresentation of female athletes, with the greatest disparity observed in 2024. The red trend line illustrates a declining trajectory in female participant inclusion across years, highlighting the persistence of gender imbalance in concussion research samples.

**Figure 3.**
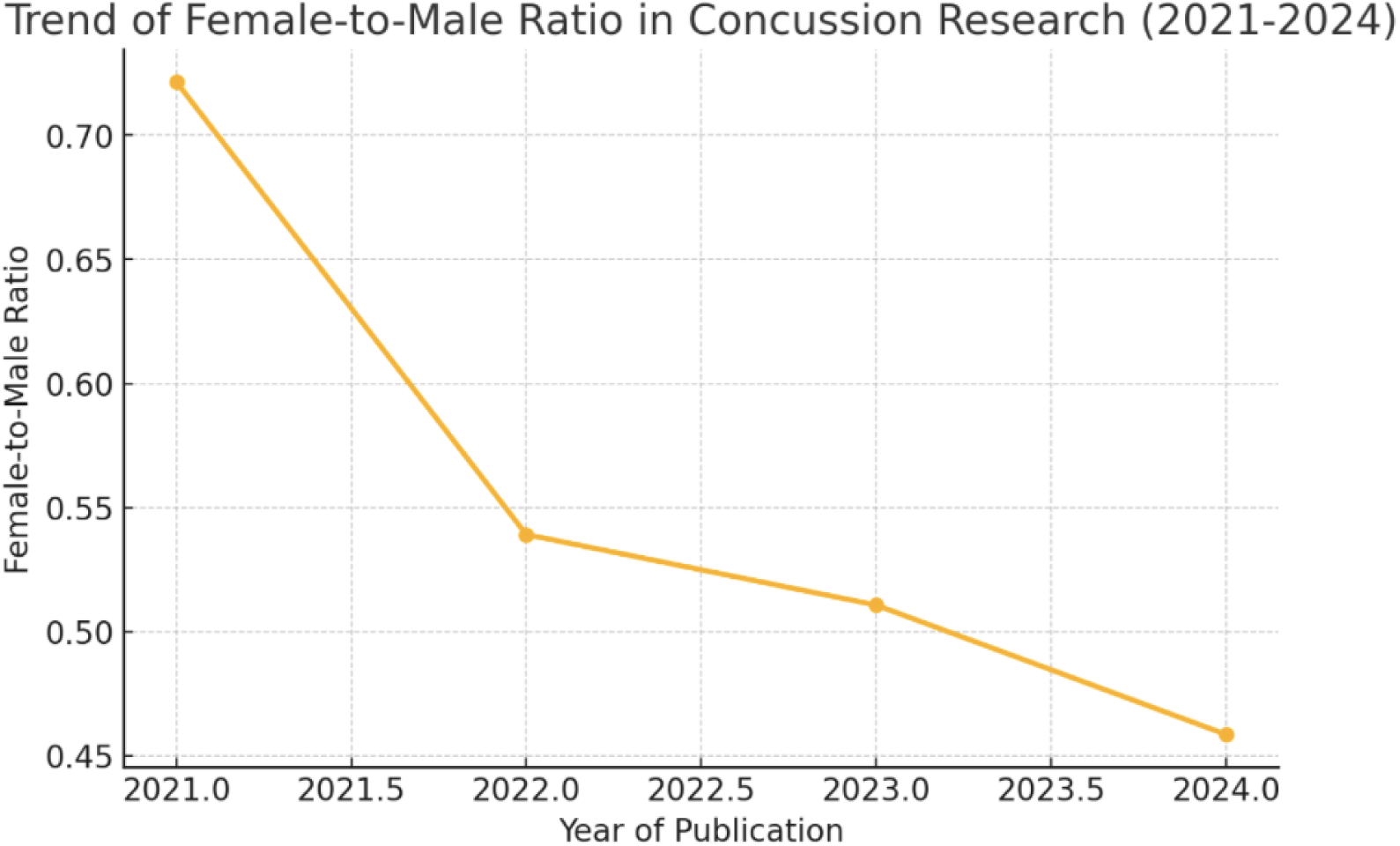
Logistic Model Regression Line: Declining female-to-male ratio in concussion research publications from 2021 to 2024. The line plot illustrates a consistent downward trend in the proportion of female participants relative to male participants across published studies, with the ratio falling from approximately 0.73 in 2021 to 0.46 in 2024.

**Figure 4.**
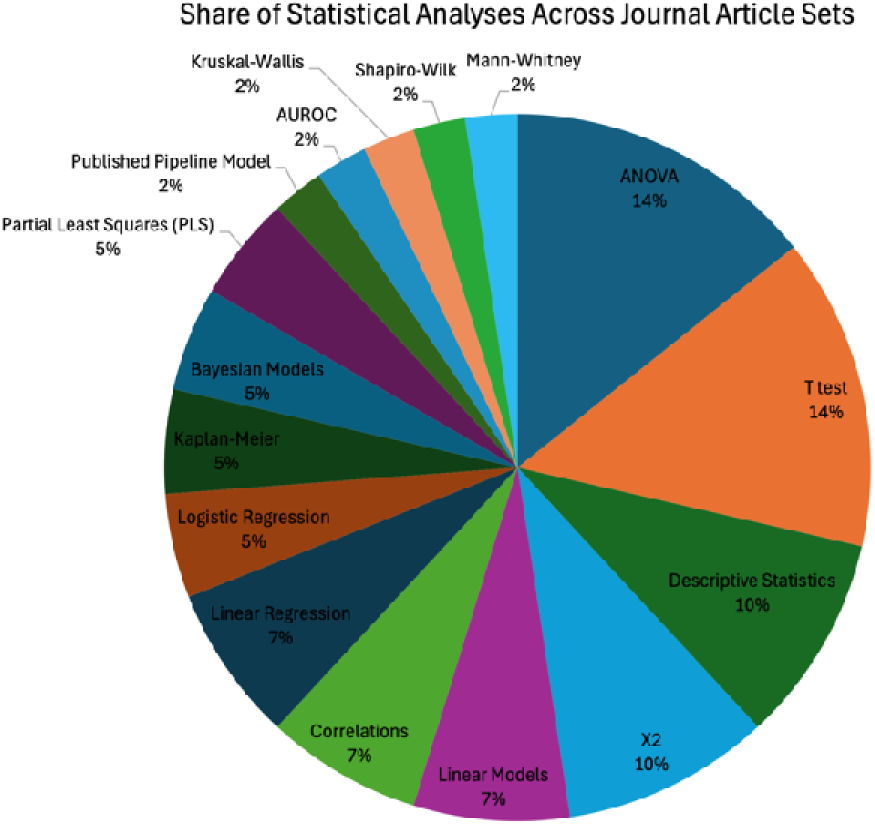
Pie Chart of Different Statistical Analyses: Chart identifying the proportion of different statistical methods used in each of the studies. Proportion of statistical methods used across concussion research studies. The pie chart illustrates the distribution of statistical analyses applied in reviewed journal articles. Traditional methods such as ANOVA and t-tests were the most frequently employed (each accounting for 14% of use), followed by descriptive statistics and chi-square tests (10% each). More advanced techniques such as logistic regression, Bayesian models, and partial least squares (PLS) regression were used less frequently, highlighting a reliance on basic analytic approaches within the current literature.

Notably, 2023 presents the largest male sample size, surpassing 600 participants, while female participant numbers remain significantly lower. The data further reveals fluctuations in female representation, with no consistent trend indicating improvement. While 2023 saw a temporary increase in female participation, the overall trend suggests a decline in female sample sizes from 2021 to 2024, with 2024 showing the lowest inclusion of female participants. This trend is particularly concerning given the growing recognition of sex-based differences in concussion outcomes. Despite increasing awareness of these disparities, research methodologies remain largely male-centered, leading to findings that may not fully apply to female athletes.

The red regression line indicates a declining trend in female sample sizes, underscoring the underrepresentation of female data in concussion research. This lack of statistically significant female sample sizes limits the validity of sex-based comparisons and hinders the development of female-specific concussion management protocols. Given the biological, biomechanical, and hormonal differences between male and female concussion responses, future studies must prioritize recruiting larger female cohorts. Without intentional efforts to close the gender gap in concussion research, clinical guidelines and return-to-play protocols will continue to be disproportionately tailored to male athletes, potentially compromising the safety and recovery of female athletes.

Next, a logistic analysis using a Poisson regression was used to examine the female-to-male ratio for each year to determine differences in sample sizes within each year. The results indicate a significant decline in female representation in concussion research over time. The analysis examined the relationship between the year of publication and female sample size, revealing a statistically significant negative association (B = - 0.2478, p <.001), suggesting that female participants have decreased despite growing awareness of sex-based differences in concussion outcomes. The model demonstrated a high explanatory power (Pseudo R² = 1.000), reinforcing the systematic underrepresentation of female athletes in concussion studies (Fig 2).

Furthermore, a female-to-male ratio analysis confirmed that male sample sizes continue to dominate, limiting the generalizability of research findings to female athletes. The analysis of female-to-male ratios across publication years reveals a declining trend in female representation in concussion research. In 2021, the female-to-male ratio was the highest at 0.72, indicating a relatively more balanced, yet still male-dominated, sample distribution. However, this ratio declined to 0.54 in 2022, 0.51 in 2023, and reached its lowest point in 2024 at 0.46. A 0.7 female-to-male ratio means that for every 1 male participant, there are 0.7 female participants in a given study. In practical terms, this indicates that female representation is only somewhat lower than male representation.

However, by 2024, if a study had 100 male participants, a 0.46 ratio would suggest approximately 46 female participants, reflecting a significant imbalance in female participation. These findings indicate that, despite growing awareness of sex-based differences in concussion outcomes, female participation in research studies has not increased proportionally and appears to be decreasing over time. This persistent gender disparity limits the applicability of concussion assessments and recovery models for female athletes, further emphasizing the urgent need for more inclusive and representative study recruitment strategies.

### Female to Male Participation Ratios

An in-depth analysis of participant demographics across various concussion studies conducted between 2021 and 2024 has revealed a troubling and consistent trend: female athletes are significantly underrepresented in the research samples. For instance, in 2021, the total sample size across the studies amounted to 1,801 participants. Among these, male participants accounted for 947 individuals, which constitutes approximately 52.6% of the sample, while female participants numbered 854, making up about 47.4%.

Although this particular year approached a near balance in terms of gender representation, this figure is somewhat misleading. A deeper exploration into the data shows that the average male-to-female ratio across these studies still persists at approximately 8:3. Such an imbalance not only raises questions about the inclusivity of concussion research but also has implications for the generalizability of the findings, as they may not adequately reflect the experiences and unique needs of female athletes.

In contrast, 2022 data reflected a more pronounced gender disparity. Of the 197 total participants that year, 128 (65%) were male and only 69 (35%) were female, resulting in a male-to-female ratio of 2:1. This trend persisted and even intensified in 2023. Among 1,743 participants, 1,066 were male (61%) and 677 were female (39%), leading to a 4:1 male-to-female ratio overall. The disparity was most pronounced in 2024, where male participants made up 214 of the 298 total sample (72%), compared to just 84 females (28%). The resulting male-to-female ratio of 6:1 indicates a substantial imbalance in representation.

The findings highlight a consistent issue of male overrepresentation in concussion research, indicating a major concern in the field. This bias may restrict the applicability of research outcomes, potentially obstructing the formulation of clinical advice tailored to different sexes. As research increasingly recognizes sex-based differences in concussion outcomes and recovery paths, addressing this disparity becomes paramount. Doing so not only guarantees fair representation in scientific research but also fosters equity in healthcare, ultimately enhancing patient outcomes for all individuals affected by concussions. Future studies must adopt a more balanced participant selection method to correct this inequality and further the understanding of concussions for both men and women.

### Effect Sizes in Research

To further evaluate the adequacy of female inclusion in concussion research, a power analysis was conducted to determine whether existing sample sizes were sufficient to detect statistically meaningful sex-based differences. Thus, a power analysis was conducted to assess the adequacy of female representation in concussion research studies and to determine whether sample sizes were sufficient to detect meaningful differences between male and female participants. The analysis was designed to evaluate the statistical power of detecting a moderate effect size (Cohen’s d = 0.5) in concussion outcomes between the two groups.

A significance level (α) of 0.05 and a power level of 80% (β = 0.80) were established, ensuring an 80% probability of identifying a true effect if one existed within the female sample. The study design considered independent group comparisons between male and female participants, making it imperative that both groups met the minimum required sample size for statistical validity.

Based on these parameters, the analysis determined that a minimum of 64 participants per group was necessary to detect a moderate effect size with sufficient statistical power. However, a systematic review of the selected studies revealed a substantial discrepancy in female representation, with only 5 out of 15 studies (30%) meeting this critical threshold. The remaining studies demonstrated a consistent power imbalance: while 7 of the 9 remaining studies met the power requirement for the male sample group, they failed to do so for the female group. This indicates that although male participant data were often robust enough to detect statistically meaningful effects, the underrepresentation of female athletes rendered findings less reliable and potentially underpowered for drawing conclusions about sex-based differences in concussion outcomes.

The implications of these findings are significant. A lack of statistical power in female samples increases the risk of Type II errors, meaning that fundamental differences in concussion symptoms and recovery patterns between male and female athletes may go undetected. Consequently, sex-specific differences in concussion pathology, recovery duration, and symptom burden may be overlooked or underestimated, reinforcing the male-dominant paradigm in concussion research. To address these limitations, future studies must ensure equitable sample sizes between sexes and implement targeted recruitment strategies to enhance female participation.

Furthermore, statistical approaches such as Bayesian hierarchical modeling or bootstrapping methods could help mitigate power imbalances in underrepresented populations, allowing for more reliable sex-specific inferences in concussion research.

### Statistical Methods in Research

The final demographic variable analyzed in this comprehensive review was the statistical methods employed across various studies. Each study uniquely utilized a diverse range of statistical techniques, which contributed significantly to the methodological diversity observed in the research. Among these methods, the analysis of variance (ANOVA) was most frequently employed, employed in six distinct studies. ANOVA is a statistical method that examines the differences in means among various groups, particularly between control and participant groups. It allows researchers to discern the significant effects of independent variables on dependent variables.

Six studies employed ANOVA and t-tests to determine statistical significance between groups. T-tests are valuable for assessing whether the means of two groups are statistically different from each other, thus providing essential insights into population differences. Furthermore, four studies relied on general descriptive statistics, which summarized and clarified key variables within those studies. Descriptive statistics provide a foundational overview of data characteristics, including central tendency and variability measures.

Another four studies applied chi-square tests, a valuable statistical technique to compare observed outcomes against expected outcomes, particularly in categorical data analysis. This method helps researchers understand the relationships between categorical variables and assess whether distributions of variables differ.

Moreover, three studies employed various linear models, including multiple linear regressions and correlation metrics, combined in unique ways to analyze the relationships between variables. Linear models are essential for examining independent variables’ linear relationships and predictive capabilities on dependent ones. In addition to the approaches above, two studies utilized more complex techniques including logistic regression, Kaplan-Meier survival analysis, Bayesian models, and partial least squares (PLS) regression. These advanced methodologies enable researchers to manage and analyze data that may not fit traditional linear models and often provide deeper insights into survival rates and probability outcomes. In conclusion, a variety of innovative techniques were employed across numerous studies to enhance the depth and rigor of the research findings.

These methods included advanced statistical analyses such as the Published Pipeline Model, which serves as a framework for organizing and evaluating research projects systematically. The studies also utilized the Area Under the Receiver Operating Characteristic curve (AUROC), a crucial measure for assessing the performance of diagnostic tests and predictive models in distinguishing between different outcomes.

Additionally, several non-parametric tests were incorporated, including the Kruskal-Wallis test, which assesses whether there are statistically significant differences between three or more independent groups. The Shapiro-Wilk test was implemented to test the normality of data distributions, a fundamental step in validating the assumptions behind many statistical techniques. Furthermore, the Mann-Whitney U test was employed to compare differences between two independent groups when the data did not meet normality assumptions.

Lastly, Fisher’s Exact Test was utilized, particularly in smaller sample sizes or when the data involved proportions, to determine if there are nonrandom associations between two categorical variables. The diversity of these statistical methods underscores the complexity and multifaceted nature of the research being conducted, highlighting the careful considerations taken to ensure robust and reliable results.

### Limitations in Statistical Modeling Due to Insufficient Female-Specific Data

While a wide range of statistical methods were employed across the studies included in this review— ranging from descriptive statistics and ANOVA to more advanced techniques such as logistic regression and Bayesian models—the vast majority of studies relied on basic, traditional analytic approaches. Importantly, this trend toward conventional statistical techniques may not simply reflect methodological preference or analytic conservatism, but rather a direct consequence of a deeper issue: the lack of robust, sex-disaggregated data, particularly regarding female athletes.

The underrepresentation of female participants limits the statistical power and variability required to support more sophisticated, multivariate, or non-linear modeling approaches. Advanced methods such as mixed-effects modeling, machine learning algorithms, and latent class analysis often require sufficiently large, well-balanced datasets to produce stable and interpretable results. In the absence of adequately sized female subgroups, researchers may be forced to collapse across sex or apply simplified models, thus perpetuating a cycle where nuanced sex-based differences in recovery trajectories go undetected.

Moreover, when female data is sparse or inconsistently collected, it becomes increasingly difficult to conduct stratified analyses or build interaction models that could account for sex-specific predictors such as hormonal status, symptom presentation patterns, or psychosocial moderators. Consequently, this lack of representation inhibits the development of predictive models that are both generalizable and inclusive, undermining efforts to individualize concussion care.

The reliance on group mean comparisons, t-tests, and ANOVA, while informative in controlled or well-balanced settings, often fails to capture the complex, dynamic, and nonlinear nature of concussion recovery— particularly among populations, like female athletes, who may follow distinct recovery patterns. As such, the continued use of basic models not only reflects the limitations of the available data but also serves as a barrier to progressive, data-driven concussion science that could inform sex-specific return-to-play protocols and personalized interventions. Addressing this methodological gap will require a systemic shift in research design, emphasizing the deliberate inclusion of female athletes, the collection of longitudinal and symptom-specific data, and the commitment to using statistical models that match the complexity of the clinical realities they are intended to represent.

## Discussion

This systematic review, along with previously conducted research, highlights the significant underrepresentation of female participants in concussion studies, which has resulted in a limited understanding of sex-based differences in recovery trajectories. The consequences of this research gap are profound, as many standardized concussion assessment tools, such as the Sports Concussion Assessment Tool (SCAT), were primarily developed using samples that predominantly included male athletes (Chin et al., 2016; Edelstein et al., 2025; Wilmoth et al., 2020). Consequently, this creates a bias that significantly reduces the applicability of these tools to female athletes and potentially compromises their safety and recovery outcomes (Covassin et al., 2013).

Furthermore, the guidelines for return-to-play (RTP) decisions are often based solely on the resolution of symptoms, which may not adequately reflect the neurobiological recovery processes occurring in females (McCrea, Broglio, & McAllister, 2021). This simplistic, one-size-fits-all approach to concussion management fails to consider the unique challenges that female athletes face. These challenges include various factors such as hormonal influences that can affect recovery rates, an increased risk of developing post-concussion syndrome (PCS), and often longer timelines for safely returning to play (Bretzin et al., 2022; Mavroudis et al., 2022; Merritt et al., 2015).

Despite a growing recognition of sex-based differences in concussion outcomes, this review, consistent with previous studies, found that simple statistical methods continue to prevail in concussion research. This reliance on basic statistical techniques limits the ability to fully understand the complex nature of concussion recovery in female athletes. Many existing studies depend on straightforward methodologies such as group mean comparisons, t-tests, or basic regression models, all of which tend to oversimplify the recovery process (CARE Consortium Investigators et al., 2018). These conventional approaches often overlook the significant individual variability in symptom resolution (Covassin et al., 2013; Master et al., 2021). Such methods typically presume a linear recovery trajectory, in which symptoms gradually improve over time. However, the reality of concussion recovery is often much more complex and nonlinear, characterized by various fluctuations influenced by individual characteristics, hormonal changes, and biomechanical factors (Meier et al., 2021).

Thus, adopting advanced statistical methodologies could pave the way for a more accurate and individualized approach to concussion assessment. Innovative machine-learning techniques, such as random forests, neural networks, and support vector machines, have the potential to identify intricate interactions among multiple variables and predict concussion recovery trajectories with significantly improved precision.

Moreover, latent class analysis (LCA) effectively classifies concussion symptom profiles into subgroups. This method allows for better identification of high-risk individuals at greater risk for prolonged recovery.

Additionally, mixed-effects modeling presents an opportunity to account for individual variability and longitudinal changes in symptoms, thereby providing a more robust framework for analyzing repeated measures data in concussion studies.

In light of these findings, there is a need to address the persistent underrepresentation of female athletes in concussion research and to refine the methodologies used to study sex-based differences in recovery.

Standardized concussion assessment tools and RTP guidelines must be adapted to reflect the unique recovery trajectories of female athletes, integrating a more comprehensive understanding of hormonal, biomechanical, and individual variability factors. Moreover, the adoption of advanced statistical techniques and machine learning approaches holds great promise for improving the accuracy of concussion assessments and personalizing treatment strategies. By prioritizing these advancements, future research can contribute to developing more equitable and effective concussion management protocols, ultimately enhancing the safety and long-term neurological health of all athletes.

### Future Directions

This systematic review, alongside a growing body of literature, underscores the ongoing and significant underrepresentation of female athletes in concussion research. As a result, the field continues to lack a nuanced understanding of sex-based differences in concussion recovery trajectories (Snook, Schaefer, & Bazarian, 2023). The implications of this gap are far-reaching. Standardized concussion assessment tools, such as the Sports Concussion Assessment Tool (SCAT), were largely developed using male-dominated samples (Chin et al., 2016; Edelstein et al., 2025; Wilmoth et al., 2020), limiting their generalizability to female populations. This male-centric design bias may compromise the validity and safety of clinical assessments when applied to female athletes, leading to inadequate care and increased risk of adverse outcomes (Covassin et al., 2013).

Further, current return-to-play (RTP) guidelines typically rely on symptom resolution as the primary criterion for recovery, neglecting the underlying neurobiological processes that may differ across sexes (McCrea, Broglio, & McAllister, 2021). This generalized approach does not account for the specific challenges female athletes face, including the influence of hormonal fluctuations, higher rates of post-concussion syndrome (PCS), and extended recovery timelines (Bretzin et al., 2022; Merritt et al., 2015). Without targeted consideration of these factors, RTP decisions may inadvertently place female athletes at higher risk for reinjury or prolonged impairment.

Despite increased awareness of sex-based disparities, most concussion studies continue to rely on basic statistical methods—such as group mean comparisons, t-tests, and linear regressions—that often oversimplify the inherently complex nature of concussion recovery (CARE Consortium Investigators et al., 2018). These techniques typically assume uniform, linear symptom resolution, failing to capture the fluctuating, individualized, and often nonlinear recovery trajectories observed in female athletes (Covassin et al., 2013; Master et al., 2021; Meier et al., 2021).

To address these limitations, future research should prioritize the adoption of advanced analytic techniques that better reflect the complexity of concussion recovery. Machine learning approaches, including random forests, neural networks, and support vector machines, offer promising avenues for modeling the dynamic interplay of biological, psychological, and environmental factors influencing recovery. Additionally, latent class analysis (LCA) can be used to identify distinct symptom profiles and subgroup recovery patterns, enhancing the ability to identify individuals at greater risk for prolonged recovery (McCrea et al., 2021). Mixed-effects models provide another robust framework for accounting for individual variability and longitudinal changes in symptomatolog, allowing researchers to model real-world recovery more accurately.

In light of these insights, future studies must deliberately include and analyze sex as a biological and social variable, ensuring that female athletes are adequately represented in both sample populations and analytic models. Concussion assessment tools and RTP protocols should be revised to integrate sex-specific risk factors, hormonal considerations, and personalized symptom trajectories. By incorporating advanced statistical methods and machine learning techniques, researchers and clinicians can move toward more equitable and data-driven approaches to concussion management. Ultimately, these efforts will support safer return-to-play decisions, improve clinical outcomes, and promote neurological health equity across all athletic populations.

## Limitations

While this review provides a comprehensive examination of recent trends in concussion research as they pertain to female athletes, several limitations must be acknowledged. First, the scope of this review was limited to studies published between 2021 and 2024, which may have excluded earlier foundational research that remains relevant to understanding long-term trends in female concussion outcomes. Additionally, while PubMed served as the primary database for sourcing studies, reliance on a single database may have inadvertently omitted relevant research published in journals not indexed in PubMed. Future studies may benefit from expanding the search to additional databases such as Web of Science, Scopus, and PsycINFO to ensure a more exhaustive review of available literature.

Second, although this review aimed to analyze the statistical methodologies used in concussion research, the classification and interpretation of statistical methods were dependent on how they were reported in each study. Some studies may have utilized advanced modeling techniques without explicitly detailing them in their methods sections, potentially leading to an underestimation of the prevalence of sophisticated statistical approaches. Similarly, the power analysis conducted in this review was based on reported sample sizes, but inconsistencies in how studies reported their data may have influenced the accuracy of these calculations.

Another key limitation is the variability in study designs and participant demographics across the studies. Differences in sample sizes, concussion severity, sport type, and age groups may have introduced heterogeneity that limits direct comparisons. Additionally, while efforts were made to ensure global representation, most studies originated from the United States, potentially limiting the generalizability of findings to other regions with different sports cultures, healthcare systems, and concussion management protocols (Covassin & Elbin, 2011; Lempke et al., 2023).

Lastly, this review primarily focused on literature investigating sex-based differences in concussion recovery and statistical methodologies used to analyze these differences. However, other critical factors, such as sociocultural influences, access to healthcare, and sport-specific policies, were not systematically analyzed.

Future research should consider a multidisciplinary approach that integrates these broader contextual factors to provide a more comprehensive understanding of the disparities in concussion research and management for female athletes.

Despite these limitations, this review highlights crucial gaps in the field. It underscores the need for more rigorous, inclusive, and methodologically advanced research to improve concussion assessment and management for female athletes.

## Conclusion

In addition to gender-equitable advancements in concussion research, there is a growing need to adopt data-driven strategies that further enhance our understanding of concussion injuries. Incorporating innovative techniques such as biomarker integration, neuroimaging analyses, and wearable technology for real-time concussion monitoring can significantly improve the precision and objectivity of concussion assessments. Researchers are better positioned to create individualized and equitable concussion management strategies by utilizing these advanced analytical techniques and proper sample sizes. This ensures that female athletes receive care tailored to their distinct recovery trajectories, ultimately improving outcomes and enhancing overall athlete health. The sporting and academic communities need to prioritize these needs and invest in a more comprehensive understanding of concussion recovery for female athletes.

## Data Availability

All data produced in the present study are available upon reasonable request to the authors

---

**Table 1.**
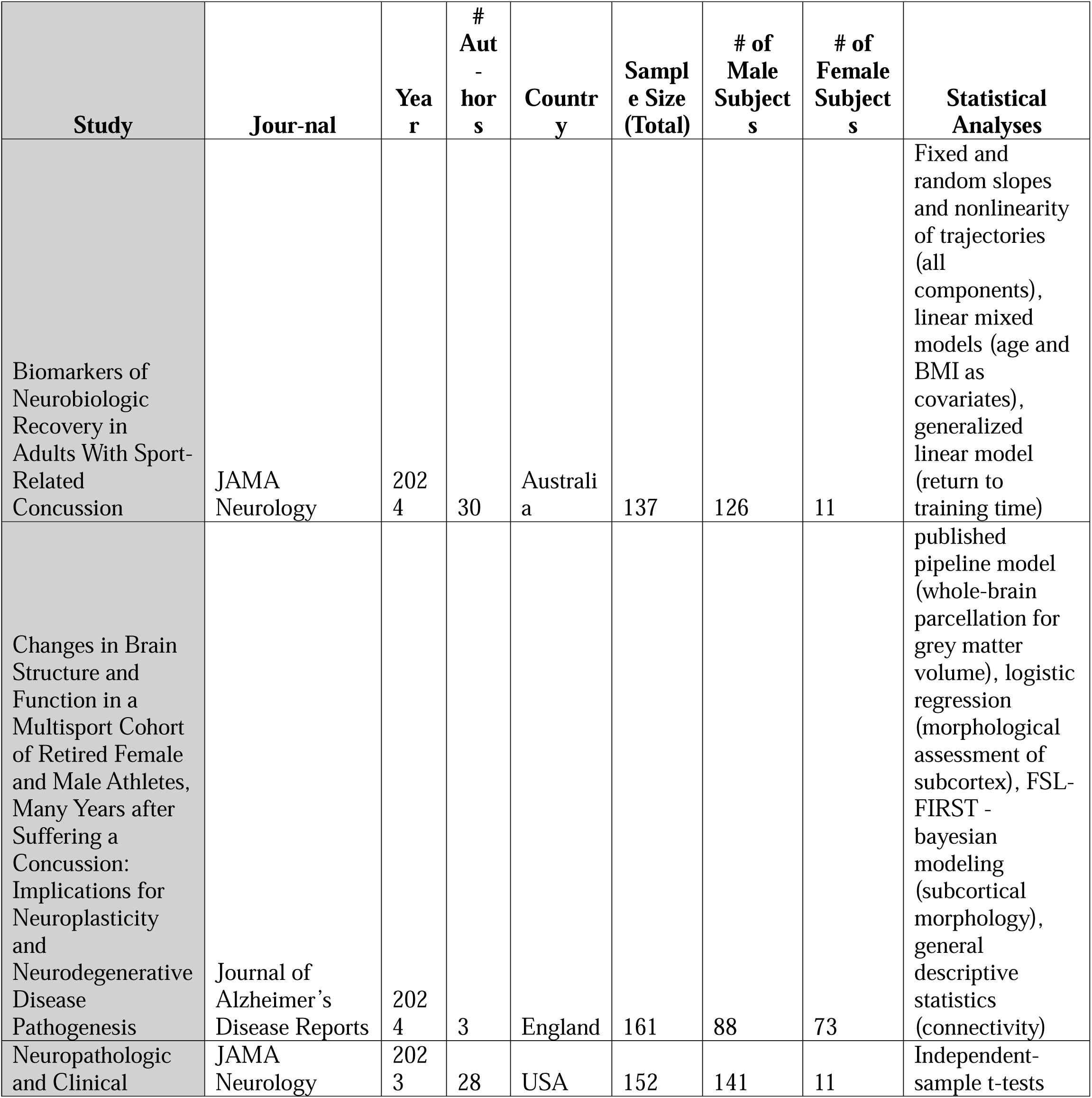
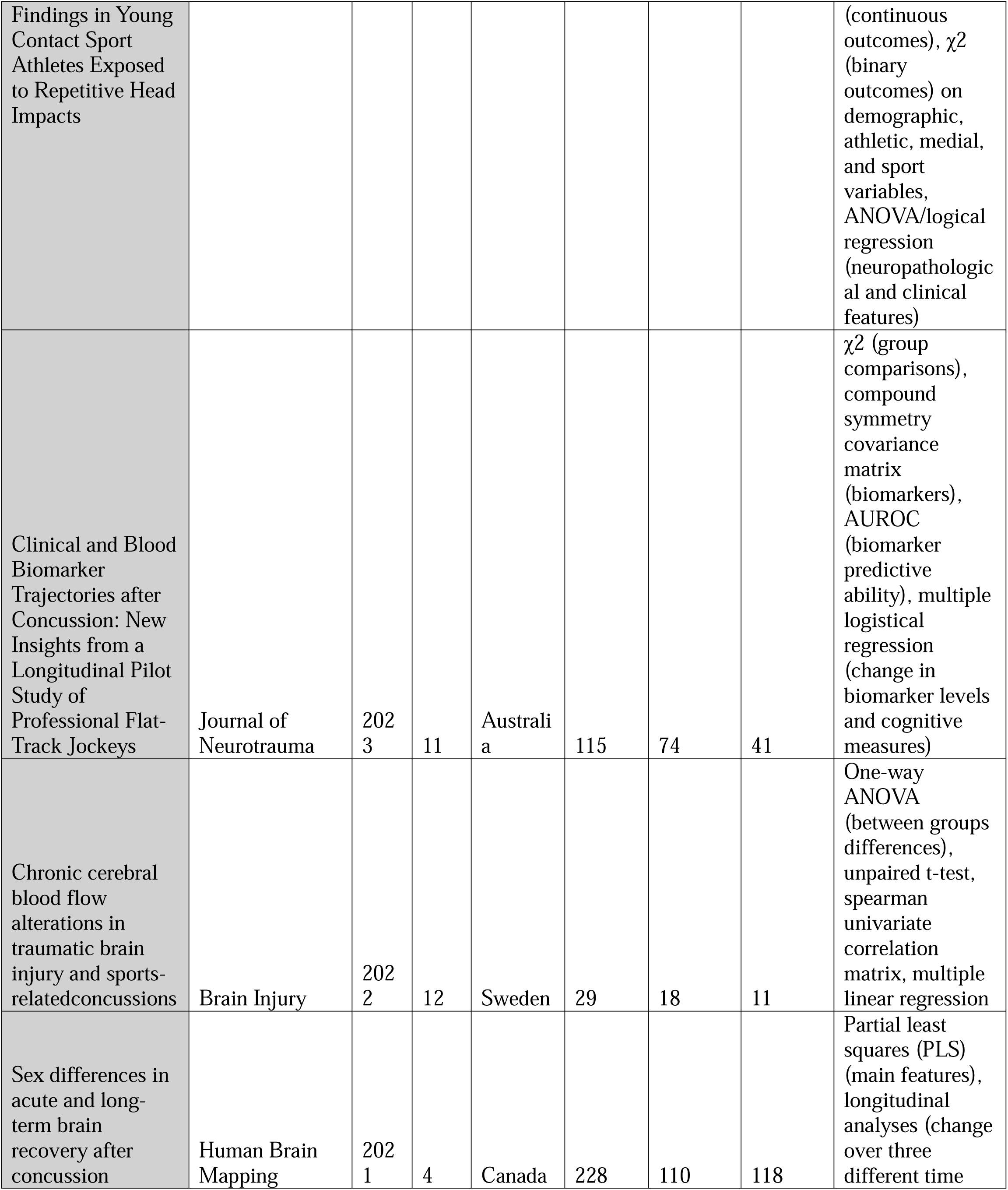
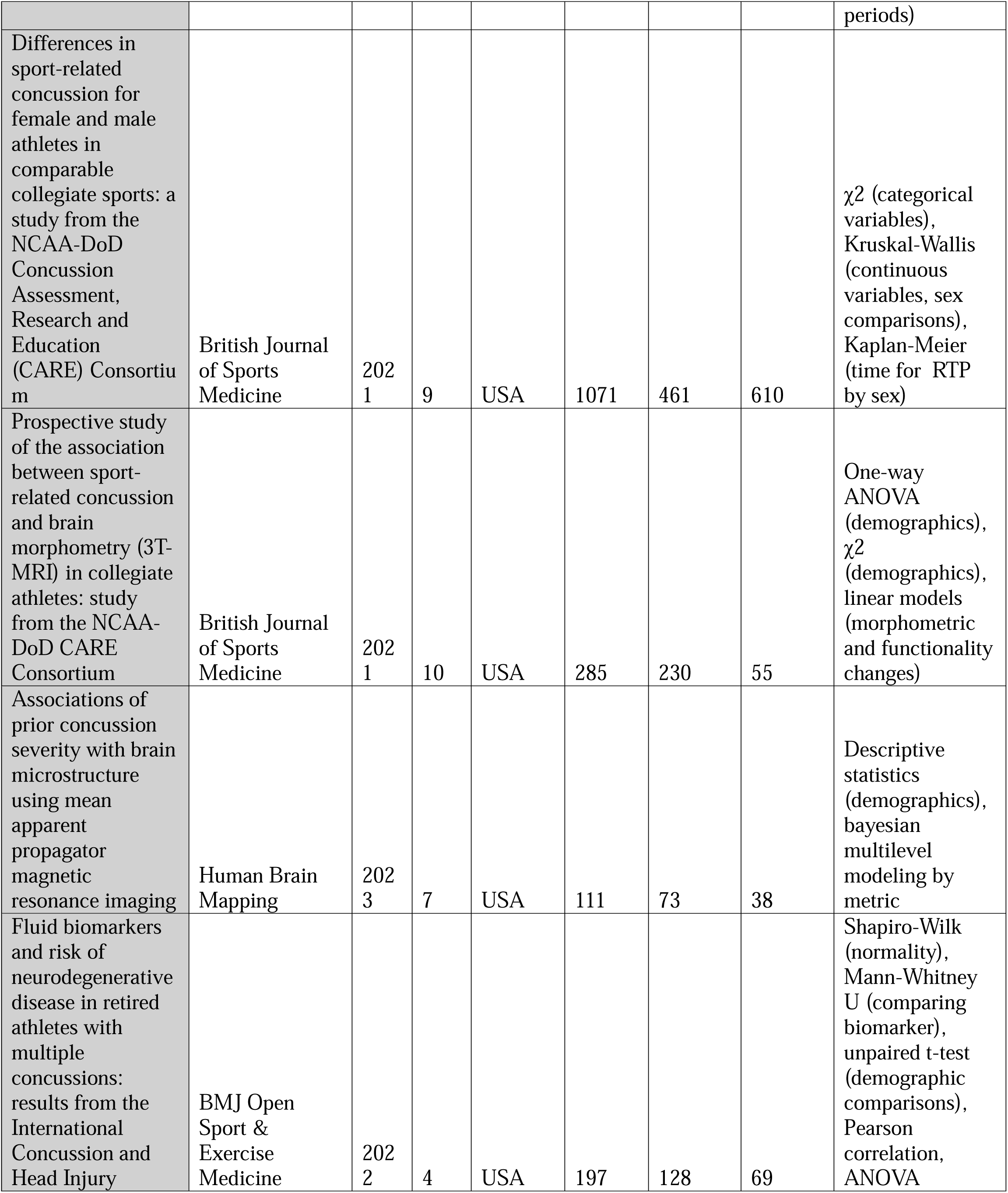
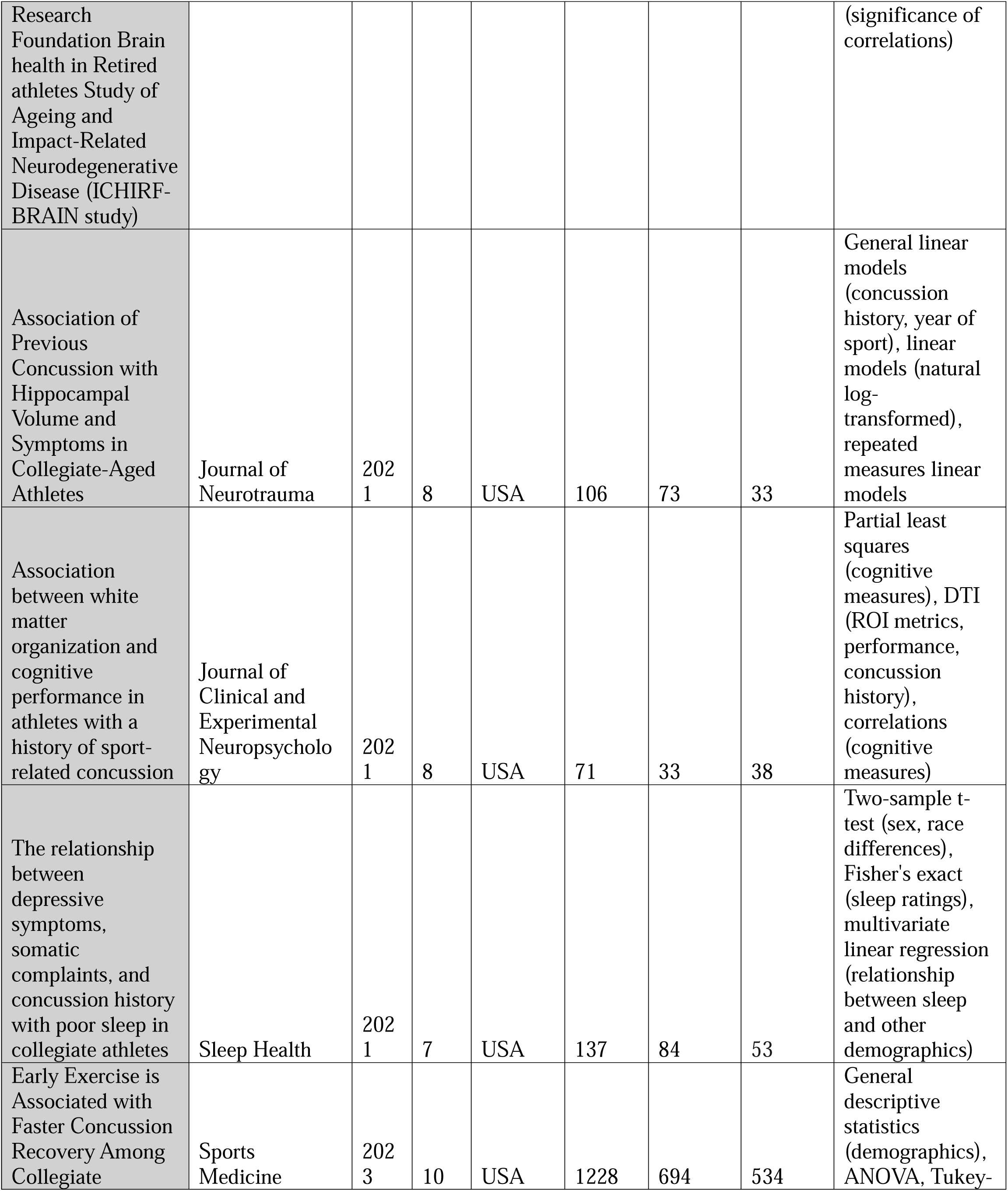
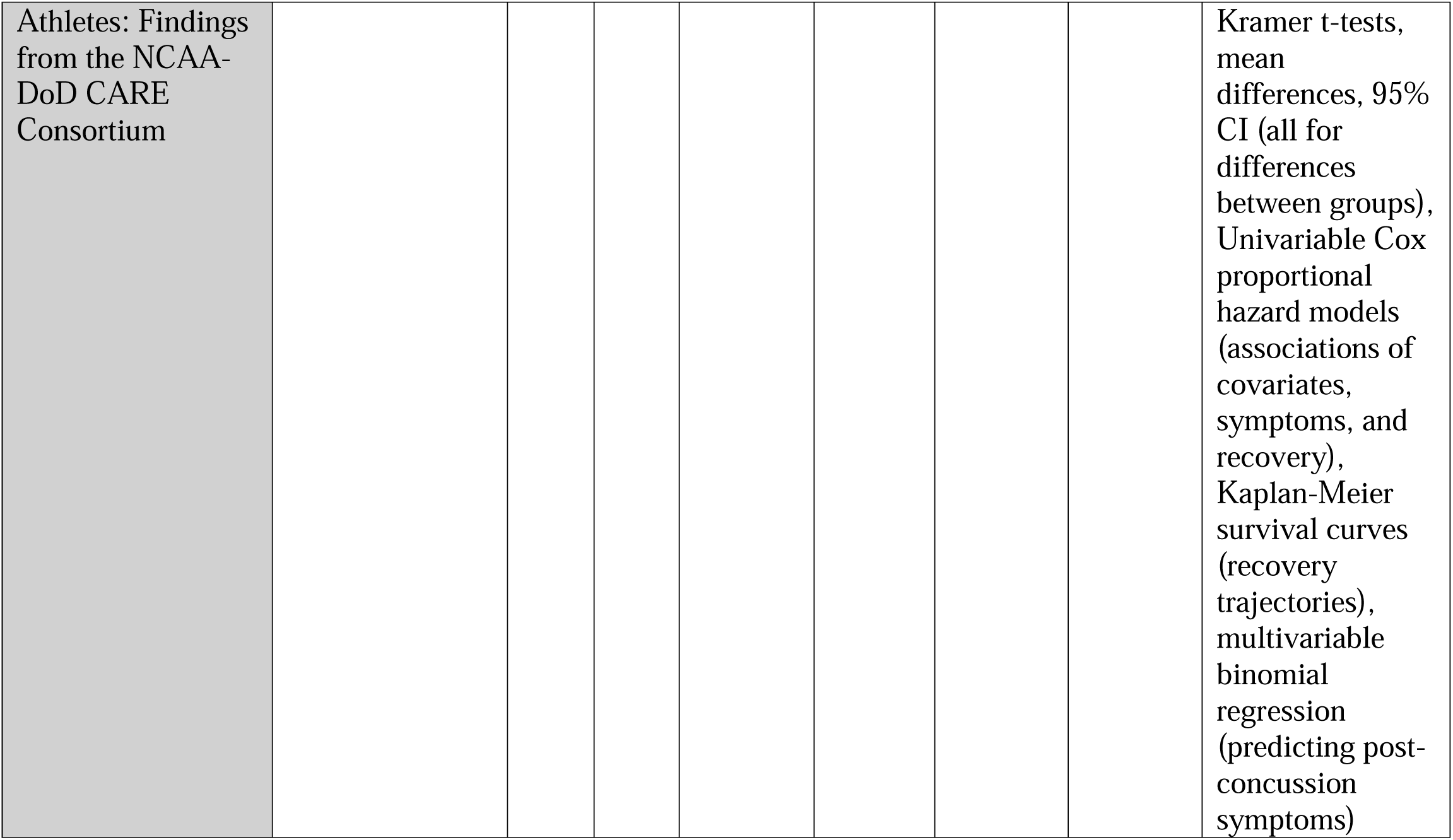
Systematic Review Articles Included: Summary of findings from the literature review on gender representation and statistical methodology in concussion research. The review synthesized data from journal articles published between 2021 and 2024, highlighting trends in participant demographics, statistical methods, and the underrepresentation of female athletes

**Table 2.** Male-to-Female Participation in Research Ratios: This table provides a detailed documentation of the overall statistics regarding male and female participation in research activities from the years 2021 to 2024. It outlines the number of participants by gender, compares trends over the specified period, and highlights any significant shifts in participation rates between male and female researchers.

| Year | Total Sample Size | Male Sample Size | Female Sample Size | Male-to-Female Ratio | Female-to-Male Ratio | Male-to-Female Ratio (X:1) |
| --- | --- | --- | --- | --- | --- | --- |
| 2021 | 1801 | 947 | 854 | 8.34 | 3.26 | 8:3 |
| 2022 | 197 | 128 | 69 | 1.86 | 0.54 | 2:1 |
| 2023 | 1743 | 1066 | 677 | 3.88 | 0.51 | 4:1 |
| 2024 | 298 | 214 | 84 | 6.33 | 0.51 | 6:1 |

